# Mapping Chemical-Gene Interactions for Developmental Lethality and Pregnancy Loss

**DOI:** 10.1101/2025.10.03.25337209

**Authors:** Syed Hassan Bukhari, Amrita Nagasuri, Boris Oskotsky, Leen Arnaout, Polina Minkovski, Gonçalo D.S. Correia, David A. MacIntyre, Gary M. Shaw, David K. Stevenson, Ruth B. Lathi, Svetlana A. Yatsenko, Tomiko T. Oskotsky, Aleksandar Rajkovic, Marina Sirota

## Abstract

Pregnancy loss affects 10–15% of clinically recognized pregnancies and arises from both genetic and environmental influences, yet evidence linking chemicals to pregnancy loss genes is rarely organized by gestational timing, maternal–fetal compartment, or lethality context. To address this gap, we integrated lethality-associated gene annotations from Intolerome with chemical–gene interactions from the Comparative Toxicogenomics Database, generating a curated network of 928 genes and ∼4,000 chemicals and developing the Chemical–Gene Atlas (CGA) to support hypothesis generation and translational research. In using recurrent pregnancy loss as a test case, the platform identified five clinically relevant genes (*F5*, *F2*, *AURKB*, *PADI6*, and *FOXD1*) with distinct exposure patterns to bisphenol A and benzo[a]pyrene, implicating gene regulation, coagulation pathways, and placental function. These findings highlight environmentally susceptible developmental windows and provide a scalable framework for investigating chemical–gene mechanisms underlying developmental lethality. The CGA is publicly available as an interactive resource to support hypothesis generation and translational research.

**Highlights:**

- Built the first interactive developmental lethality chemical-gene atlas for prenatal and postnatal disease research.
- Integrated 4,110 chemicals with 928 developmental lethality-associated genes into a unified exposomic framework.
- Identified vulnerable fetal and second-trimester developmental windows through timing and maternal-fetal annotations.
- Identified convergent interactions between BPA, B[a]P, and five key RPL-associated genes (F5, F2, AURKB, PADI6, and FOXD1), implicating coagulation, placental, and embryonic development pathways in pregnancy loss.

## Introduction

Pregnancy loss, specifically miscarriage before 20 weeks of gestation, affects 10-15% of all clinically recognized pregnancies, and rates are much higher if preclinical and biochemical pregnancy losses are included.^1^ Recurrent pregnancy loss (RPL), defined as two or more pregnancy losses, affects ∼5% of couples.^2^ RPL takes an emotional and psychological toll on families and individuals, leading to anxiety and uncertainty about future pregnancies.^3^ Understanding the genetic etiology and environmental influences of RPL can inform family planning and targeted interventions, as research increasingly demonstrates that these factors impact embryonic survival and pregnancy outcomes.^4^

Environmental exposures further compound this risk. Endocrine-disrupting chemicals (EDCs) interfere with the body’s hormonal balance and can impact fertility and pregnancy outcomes.^4,5^ Heavy metals such as lead, mercury, and cadmium can also result in fetal growth restriction, neurodevelopmental complications, and placental insufficiency.^6^ Air pollutants, such as particulate matter, nitrogen dioxide, and carbon monoxide, can reduce placental blood flow, increase inflammatory and oxidative stress responses that damage fetal DNA, and increase the risk of preterm birth or even stillbirth.^7^

We hypothesized that genes essential for human embryonic development and susceptible to chemical perturbation represent priority candidates for mediating exposure-driven pregnancy loss, particularly when gene functions or pathways are vulnerable to disruption by multiple chemical exposures.

Gene-environment interactions (G × E), including chemical-gene interactions, are central to developmental disease risk. However, current toxicogenomic resources are poorly aligned with pregnancy-loss decision-making. They typically lack annotations for gestational timing, maternal-fetal compartment, and lethality, and they do not systematically prioritize high-risk developmental genes. As a result, researchers cannot readily ask which exposures matter, when in gestation they matter, and in which compartment they may act to cause pregnancy loss.

For instance, the Intolerome database catalogs 934 genes essential for fetal viability, most of which reflect fetal genetic contributions, but it does not provide a structured framework for exploring environmental or chemical interactions in the exposure context.^8,9^ The Comparative Toxicogenomics Database (CTD) provides a free, peer-reviewed resource containing meticulously curated information on chemical-gene-disease interactions.^10^ While valuable, CTD does not allow exploration of chemical–gene interactions by developmental stage, affected biological systems, or lethality phases, thereby limiting its relevance for reproductive toxicology and RPL research. Its output remains largely static and tabular, restricting deeper network-level analysis. CTD also lacks integrative tools that combine gene intolerance, chemical exposure, and developmental timing.

To address this gap, we developed the Chemical-Gene Atlas (CGA), an interactive, publicly accessible exposomic resource that integrates loss-of-function intolerant gene data from the Intolerome with curated chemical-gene interaction data from the CTD. The CGA enables users to filter and visualize ∼900 genes and 4,000 chemicals by developmental timing, biological system, gene effect, and functional mechanism, providing a novel means to map the interactions between environmental exposures and genetic vulnerability. CGA represents the first effort to systematically unify intolerance, exposure, and developmental context into an interactive platform for research on pregnancy loss. In this framework, an “interaction” is an environmentally induced perturbation mapped onto a loss-of-function–intolerant gene, contextualized by the gene’s effect, developmental timing, the system affected, and the lethality mode.

Our approach integrates genetic intolerance information from the Intolerome database with chemical-gene interactions reported in the CTD and specific to distinct developmental outcomes. CGA does not replace existing curated resources or introduce a new primary source of chemical-gene evidence. Rather, it adds value by integrating fragmented toxicogenomic and developmental-lethality annotations into a single pregnancy-loss-focused framework. This allows users to explore chemical-gene relationships alongside developmental timing, affected system, gene effect, and lethality context in a single interface, making cross-resource evidence more interpretable and actionable for hypothesis generation in reproductive and developmental disease research. As an illustrative use case, we applied CGA to recurrent pregnancy loss (RPL) to explore how environmental exposures interact with fetal gene networks.

## Results

### Study design

Our study integrated two large-scale datasets, Intolerome and CTD, to build an exposomic resource focused on chemical-gene interactions associated with developmental lethality (Figure 1). As summarized in Figure 1, the Intolerome dataset comprised 934 genes categorized as prenatal and postnatal genetic intolerance, which were used to identify suitable chemicals. Subsequently, the interactions involving these 934 genes in CTD focused on *Homo sapiens*. This merge reduced the ∼10,000 unique chemicals and about 1.2 million chemical-gene interactions in CTD to 928 genes, ∼4000 unique chemicals, and ∼73000 total interactions (Figure 1). By restricting CTD interactions to those involving only the 928 intolerome genes in *Homo sapiens*, we provide a robust and interpretable source for exposomic network analysis, pathway enrichment, and the identification of vulnerable genes for various chemical exposures.

**Figure 1.**
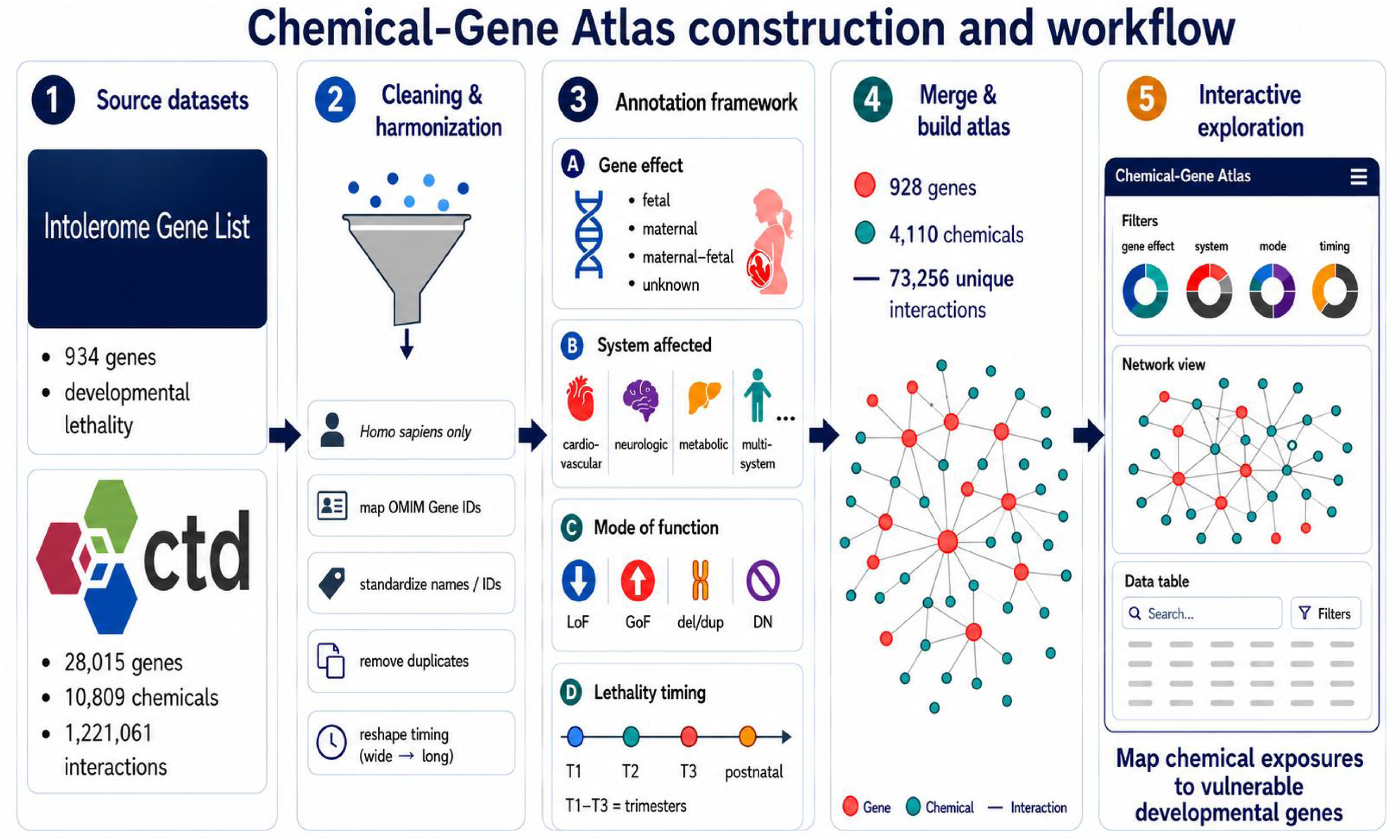
Construction of the Chemical-Gene Atlas and interactive workflow. The workflow integrates an Intolerome gene list of 934 developmental lethality-associated genes with CTD (Comparative Toxicogenomics Database) records, which contain 28,015 genes, 10,809 chemicals, and 1,221,061 chemical-gene interactions. After harmonizing the data by restricting to *Homo sapiens*, mapping OMIM identifiers, standardizing gene names, removing duplicates, and restructuring timing annotations, the dataset was organized by gene effect, affected biological system, mode of function, and lethality timing across developmental stages. Merging these annotations with CTD interaction data produced the final Chemical-Gene Atlas, comprising 928 genes, 4,110 chemicals, and 73,256 unique chemical-gene interactions, which can be explored through interactive filters, network visualization, and searchable tables.

### Distribution of Gene Effects, Systemic Impacts, and Lethality Timing

After merging the dataset, we characterized the developmental relevance of the 928 genes in the merged dataset across four biological categories: gene effect (fetal vs maternal), affected physiological systems, lethality mode of function (loss of function, gain of function, etc), and timing of lethality (Figure 2). The merged dataset was used to build an interactive tool, CGA, available as an R Shiny application called “cgatlas” (https://cgatlas.org/).

**Figure 2.**
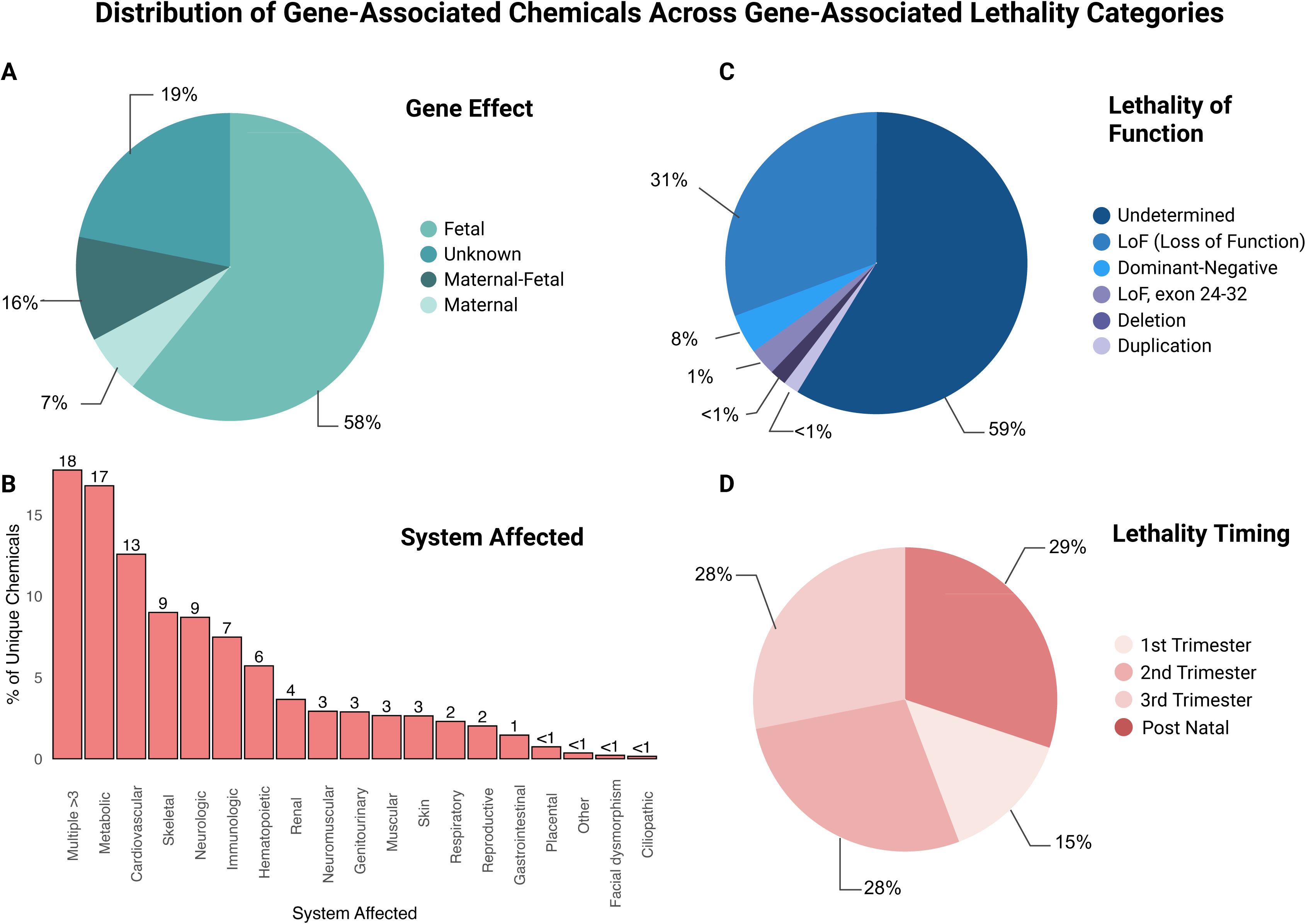
Distribution of Gene-Associated Chemicals Across Gene-Associated Lethality Categories. **(A)** Gene effect: most chemicals are associated with fetal-specific effects, with smaller subsets linked to maternal, maternal–fetal, or unknown origins in the merged dataset. **(B)** Physiological systems affected: chemicals most frequently impact the metabolic and multi-system categories, followed by cardiovascular and neurological systems. Less frequent associations occur in skeletal, immunologic, renal, and other systems. **(C)** Lethality mode of function: most associations are undetermined, with a substantial subset classified as loss-of-function (LoF). Additional categories include dominant-negative, LoF (exon 24–32), deletions, and duplications. **(D)** Lethality timing: chemicals are distributed across all trimesters, with a slightly greater contribution in postnatal lethality. Entries without defined lethality categories were excluded from this visualization.

As shown in Figure 2A, 58% of unique chemicals are associated with fetal-specific effects that directly impact embryonic or fetal development. Approximately 16% are linked to both maternal and fetal physiology. About 7% are associated with maternal-only effects, while 19% may have unknown effects.

The distribution of genes across affected biological systems is shown in Figure 2B. The multi-systemic disorders had the highest number of unique chemicals (18%), followed by the metabolic system (17%) and the cardiovascular system (13%). Other systems with notable involvement were neurologic (9%), skeletal (9%), and immunologic (7%). As shown in Figure 2C, most gene disruptions were assigned to an undetermined functional category. Among the classified variants, LoF events were the most prevalent, followed by dominant-negative effects, exon 24–32 specific LoF (FBN1), and structural variants such as deletions and duplications.

Lastly, Figure 2D shows the distribution of unique chemicals associated with lethality events across developmental timepoints: unique chemicals are most frequently implicated in postnatal outcomes (∼29%), followed by the second (∼28%), third (∼28%), and first (∼15%) trimesters. These distributions suggest that environmentally linked developmental lethality is concentrated in fetal-specific contexts and is especially prominent in multisystem, metabolic, and cardiovascular biology, thereby highlighting developmental windows and physiological systems that may be particularly vulnerable to chemical perturbation.

### Chemical-Gene Interaction of RPL

The applicability of this application can be described by providing an example of a disease and mapping its chemical-gene interactions. Therefore, we have extracted focused subnetworks of five integral genes (*F5, F2, AURKB, PADI6*, and *FOXD1*) associated with recurrent pregnancy loss (RPL), selected for their roles in embryonic development, fertility, and survival during early life stages.^2,11–12^

Each RPL gene was mapped with its complete exposomic profile, visualized as networks in Figure 3A. A chemical was considered “linked” to a gene if a curated interaction between the two was present in CTD (e.g., documented effects on expression, activity, or binding, restricted to *Homo sapiens*). Node degree, defined as the number of unique chemical exposures interacting with a given gene, serves as a quantitative proxy for environmental connectivity or sensitivity here. Using this measure, *F5* and *AURKB* genes exhibited the highest node degrees, whereas PADI6 showed a compact network with a narrower exposomic footprint.

**Figure 3.**
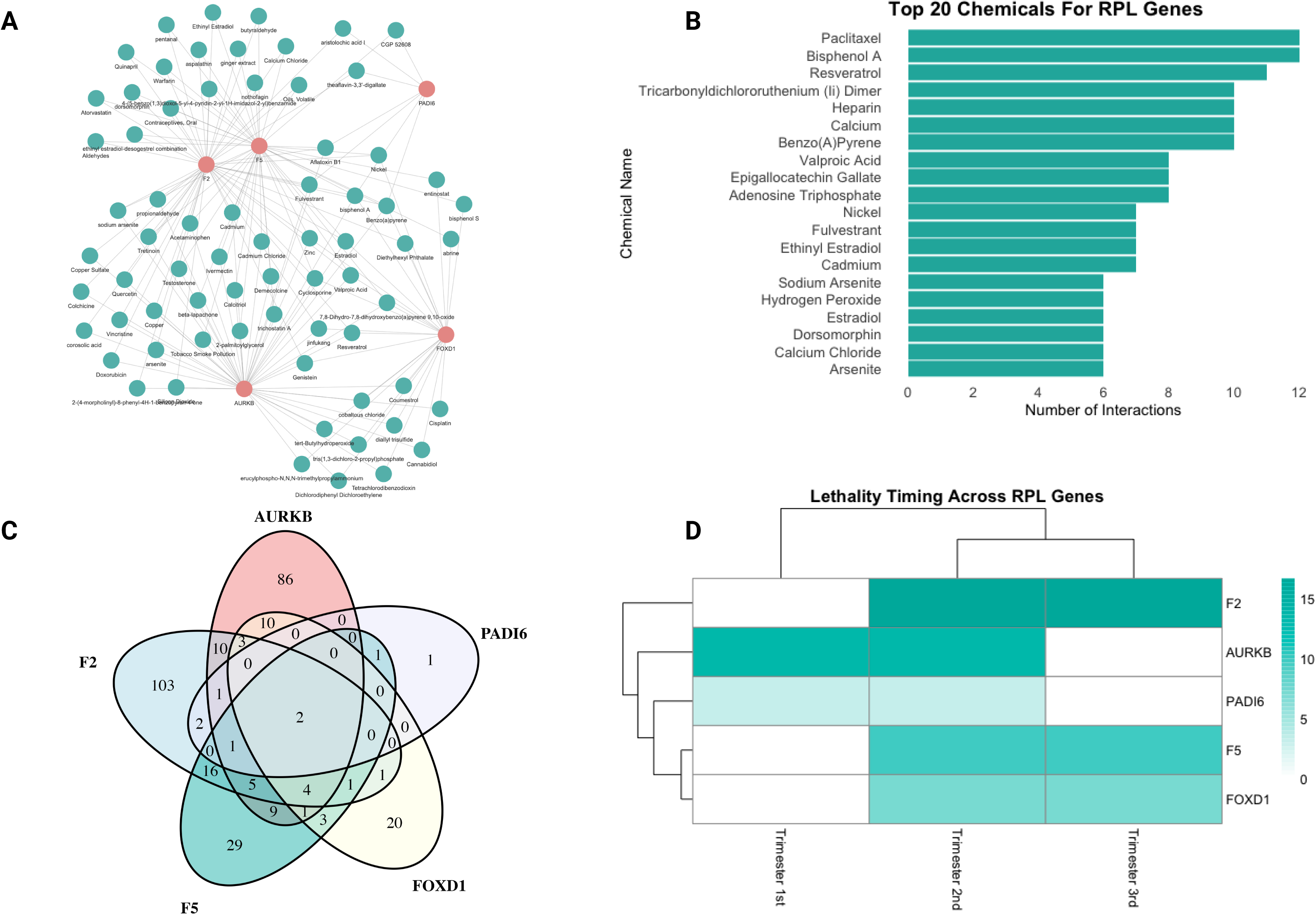
Analysis of Recurrent Pregnancy Loss (RPL) Genes, Chemicals, and Interactions. **(A)** Network visualization showing interactions between RPL genes (red nodes) and chemicals (green nodes). Each chemical node is connected to at least two RPL genes, demonstrating shared chemical associations and high interconnectivity across all five genes. **(B**) Bar plot showing the top 20 chemicals associated with RPL genes, ranked by the number of interactions. Paclitaxel, bisphenol A, and resveratrol are among the most frequently linked chemicals. **(C)** Venn diagram showing the overlap of chemicals across five key RPL-related genes (*AURKB*, *PADI6*, *F2*, *F5*, and *FOXD1*), highlighting the unique and common chemical-gene interactions among RPL genes. The center shows two main chemicals (Benzo(a)pyrene and bisphenol A) that interact with all five genes. **(D)** Heatmap of lethality timing across different trimesters for selected RPL genes, with a log scale, indicating variations in critical time windows for gene-associated lethality risks.

Additional information on the timing of lethality events is provided in Figure 3D, which shows that none of these genes affect postnatal outcomes, but all are implicated in the second trimester.

The Venn diagram and network plots were complemented by showing the top 20 most frequently associated chemicals for RPL in Figure 3B. These chemicals are ranked by their number of interactions across the five selected RPL genes to provide a high-level overview of prevalent exposures within the RPL gene set. This figure reflects bisphenol A and Benzo(a)pyrene, the two chemicals common among all five genes, as having the second and sixth highest number of interactions with RPL genes.

We further focused our analysis by comparing shared versus unique environmental risks across all the RPL genes, as shown in Figure 3C. This Venn diagram represents the overlap in chemical interactors. Although most genes displayed distinct interaction profiles, bisphenol A and Benzo(a)pyrene stood out, as they interact with all five RPL genes. In addition to overlap counts, each gene–exposure pair can be annotated with contextual features, such as node degree, the number of curated CTD interactions supporting that link, and the developmental timing of lethality, providing a prioritization signal for researchers designing mechanistic follow-up and exposure-focused studies.

Extending upon our exploration of the interactions between RPL genes and associated chemicals in Figure 3A, we have isolated the exposomic networks for the five RPL genes, shown in Figure 4. The size and density of each network reflect the underlying literature and annotation density rather than true exposure burden. The genes *F5* and *AURKB* have the highest number of connections and broad exposomic profiles. In contrast, *PADI6* displays a smaller, more focused network. Lastly, *FOXD1* and *F2* fall in between, with a moderate number of chemical connections. These differences in exposomic network structure suggest that RPL-associated genes may vary in the breadth of their environmentally linked perturbation, thereby helping distinguish broadly connected candidate genes from those with narrower but potentially more specific exposure profiles.

**Figure 4.**
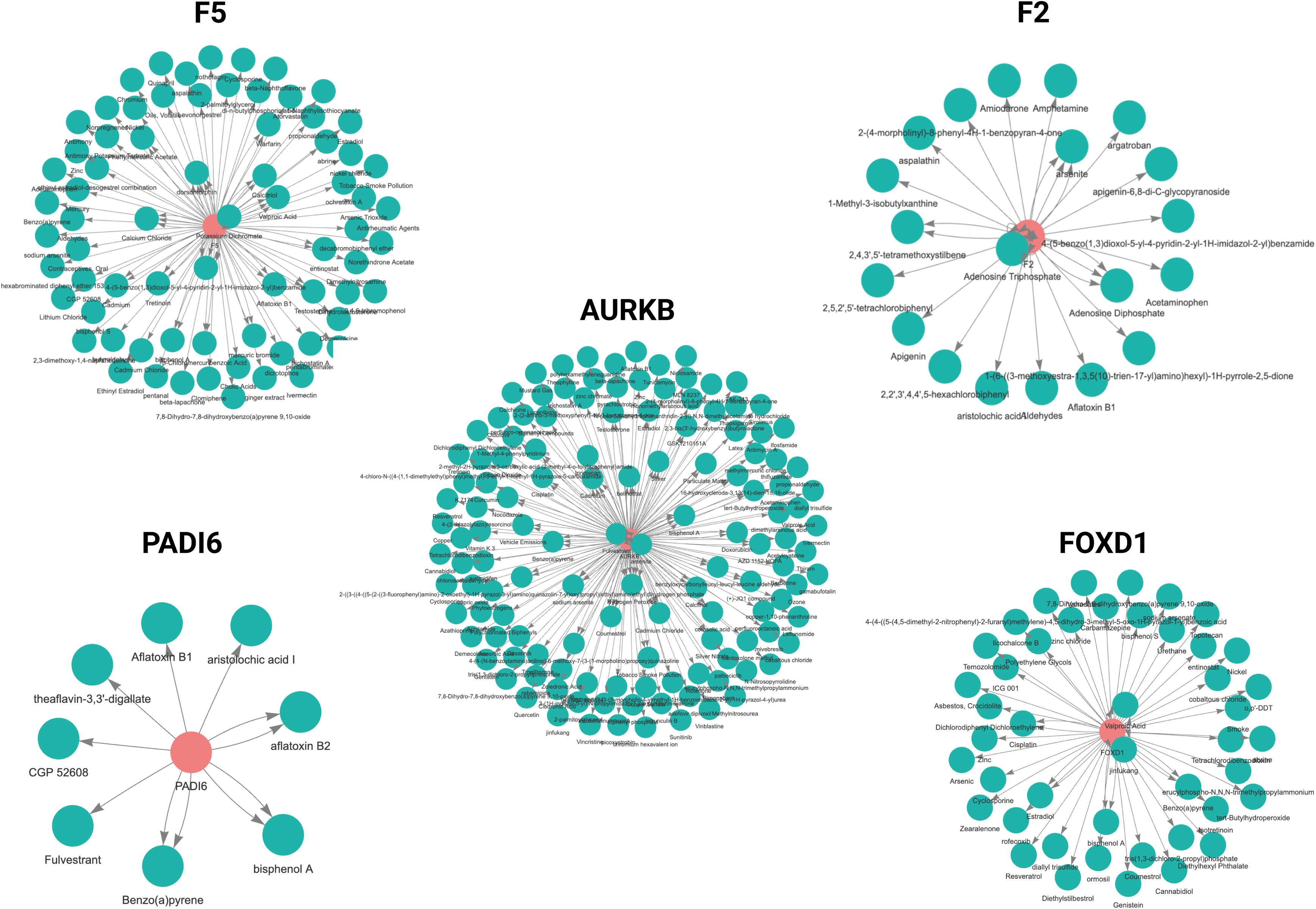
Exposomic Networks for Chemical-Gene Interactions of RPL Genes. Exposomics interaction networks show chemical associations for five genes related to RPL in genetic lethality and dysfunction. Each exposome network is centered around a gene (red node) and its chemical interactors (green nodes) derived from the Comparative Toxicogenomics Database (CTD). The networks differ in their susceptibility to environmental exposure. *F5* and *AURKB* exhibit extensive chemical interactions, indicating broad environmental sensitivity, whereas *PADI6* demonstrates limited interactions, suggesting a more specialized exposome profile. *F2* and *FOXD1* networks display intermediate connectivity. The collection of these exposome profiles relates to chemical exposures and genetic expression, disease progression, and developmental outcomes.

Figure 5A summarizes BPA-associated interactions across all five RPL genes, including increased or decreased expression for *FOXD1* and *F5*, altered methylation for *AURKB* and PADI6, and changes in protein abundance or enzymatic activity for *F2*. Figure 5B shows that benzo[a]pyrene also perturbs all five genes, with largely repressive effects, including decreased expression and methylation changes across the subnetwork.

**Figure 5.**
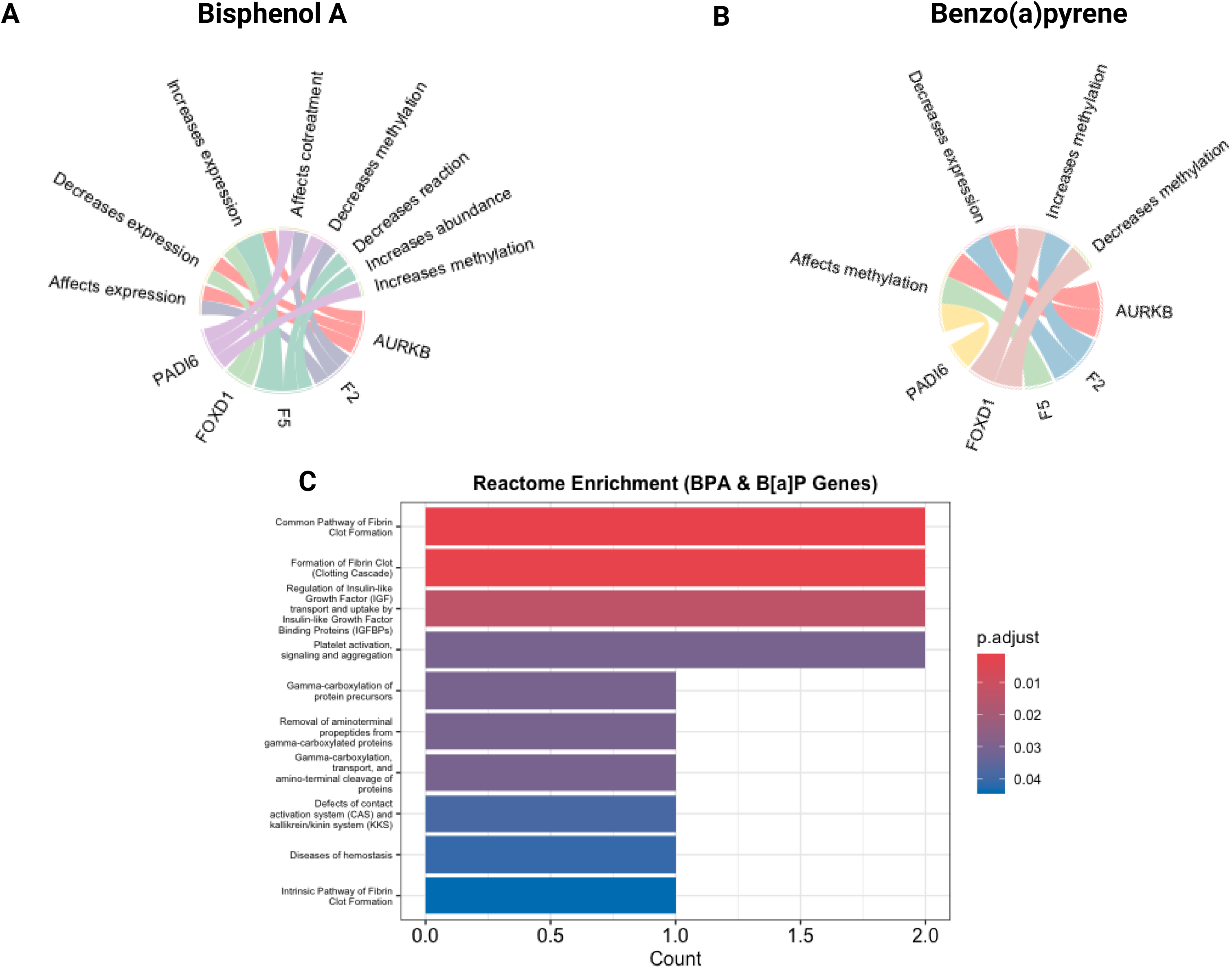
Chord diagrams and Reactome pathway enrichment for bisphenol A (BPA) and benzo[a]pyrene (B[a]P). Chord diagrams **(A, B)** show chemical–gene interactions for five RPL-associated genes (*AURKB, F2, F5, FOXD1*, and *PADI6*), with ribbons connecting each specific molecular action (e.g., “increases methylation,” “decreases expression”) to its respective gene target. Interaction data was obtained from merging the Intolerome and CTD datasets. BPA exhibited diverse regulatory effects, including increased methylation and gene expression, whereas B[a]P mainly induced repressive changes, such as decreased methylation and reduced gene expression. Barplots **(C)** present Reactome pathway enrichment analyses of genes linked to BPA or B[a]P with significant enrichment (FDR-adjusted p < 0.10) primarily in pathways related to hemostasis, including “Common Pathway of Fibrin Clot Formation,” “Formation of Fibrin Clot (Clotting Cascade),” and “Platelet Activation, Signaling, and Aggregation.” P-values were calculated using a hypergeometric test and adjusted for multiple comparisons using the Benjamini–Hochberg method.

Panel 5C provides pathway-level support for these interactions by showing enrichment of coagulation-and implantation-relevant biological processes. Both BPA and B[a]P exposures are enriched for disruptions in fibrin clot formation, coagulation cascades, and gamma-carboxylation of protein precursors. These biological pathways directly involve *F2* and *F5*.

The molecular interactions (as shown in Figure 5A and 5B) show that both BPA and B[a]P have curated CTD interactions with the same set of RPL genes most enriched in our developmental lethality dataset, while the pathway enrichment (Figure 5C) reveals a shared downstream disruption of coagulation and implantation-related biology.

### Benchmarking

To contextualize CGA relative to existing toxicogenomic, exposomic, and disease-association resources, we benchmarked its core capabilities against CTD, ToxCast/CompTox, AOP-Wiki, DisGeNET, Open Targets, and ExposomeExplorer (Table 1). This comparison was intended to clarify the specific gap CGA addresses rather than to suggest that these resources are interchangeable. Existing platforms provide important complementary strengths, including curated chemical–gene interactions, high-throughput assay data, adverse outcome pathway organization, gene–disease associations, and exposure biomarker information. Resource capabilities were qualitatively categorized based on publicly documented database functionality, annotation structure, and relevance to developmental and reproductive toxicology workflows, where “High” indicates extensive and directly integrated support for a feature, “Moderate” indicates partial or indirect support, “Low” indicates limited support, and “None” indicates the feature is not systematically represented.

**Table 1.** Comparison of CGA with related toxicogenomic, exposomic, and disease-association resources.

| Tool | Chemical-Gene Interaction | Disease Context | Developmental/ Reproductive Context | Timing Filters | Maternal/ Fetal Annotation | Network View | Assay Data | Exposure Biomarkers | References |
| --- | --- | --- | --- | --- | --- | --- | --- | --- | --- |
| **CGA** | High | High | High | High | High | High | None | None | – |
| **CTD** | High | High | Low | Low | Low | Moderate | None | Low | Davis et al.<sup>10</sup> |
| **ToxCast / CompTox** | Low | Low | Low | Low | None | Low | High | Moderate | Williams et al.<sup>13</sup>; EPA CompTox<sup>14</sup> |
| **AOP-Wiki** | Low | Moderate | Low | Low | None | Moderate | None | None | Martens et al.<sup>15</sup> |
| **DisGeNET** | None | High | Low | None | None | Low | None | None | Piñero et al.<sup>16</sup> |
| **Open Targets** | None | High | Low | None | None | Low | None | None | Buniello et al.<sup>17</sup> |
| **ExposomeExplorer** | None | Low | Low | None | None | None | None | High | Neveu et al.<sup>18</sup> |

However, most existing resources do not integrate chemical–gene interactions with developmental or reproductive disease context, maternal/fetal annotation, lethality timing, and network-level visualization in a single framework. Table 1, therefore, highlights CGA’s added value as a pregnancy-loss-focused integration layer that connects toxicogenomic evidence with developmental lethality annotations and enables more targeted exploration of environmentally linked reproductive risk.

## Discussion

Our exposomic analysis identified associations between gene intolerance, developmental lethality, and exposure to environmental chemicals. Pairing chemical-gene interaction data from the Comparative Toxicogenomics Database (CTD) with Intolerome enabled the categorization of nearly 928 genes and ∼4000 unique chemicals. Most of these genes were intolerant to loss-of-function (LoF) and were enriched in the metabolism, cardiovascular, and neurological pathways. The temporal concentration of lethality was observed during late gestation and the postnatal phase of development, indicating that variants in these genes may be present during specific developmental stages, such as during physiological stress during immune maturation and organogenesis.

The majority of the chemicals involved in developmental lethality are annotated to fetal rather than maternal pathways (Figure 2A). This trend suggests that environmental exposures are more often connected to fetal-specific mechanisms, though it may also reflect the current emphasis of available studies and curated toxicogenomic data.

In profiling five RPL-associated genes, *F5*, *F2*, *AURKB*, *PADI6*, and *FOXD1*, we identified distinct exposomic network structures (Figure 3A–D, Figure 4). The exposomic profiles showed that the *F5* and *AURKB* genes exhibit the highest node degrees, which may reflect broad environmental sensitivity, whereas *PADI6* had a compact network with a narrower exposomic footprint.

*FOXD1* and *F2* fell in between, with a moderate number of chemical connections. *FOXD1* regulates developmental patterning, while *F2* (thrombin) contributes to coagulation cascades; ultimately, both are integral to implantation and placental stability.^2,25^ Narrowing down on each gene network provides a clear comparison of how different RPL genes are environmentally loaded, with *F5* and *AURKB* representing broad exposure profiles and *PADI6* reflecting sensitivity to a more limited but potent set of exposures.

Two chemicals, bisphenol A (BPA) and benzo[a]pyrene (B[a]P), interacted with all five genes, implicating shared toxicological mechanisms. Mechanistic annotations indicate that BPA and B[a]P modulate gene expression and methylation, particularly affecting pathways involved in coagulation and placental function. BPA is a widely used endocrine disruptor in polycarbonate plastics and thermal papers, resembling estrogenic action in breaking implantation, causing placental malfunction, and distorting fetal programming.^26,27,34–36^ Within the context of known biological effects (Figure 5A), BPA exposure causes transcriptome reprogramming and altered epigenetic marking phenotypes across the placenta and embryo.^27,28^ By contrast, B[a]P, a polycyclic aromatic hydrocarbon formed by incomplete combustion, primarily acts by activating the aryl hydrocarbon receptor (AhR), leading to oxidative stress, DNA adduct formation, and dysregulation of gene expression.^29,30^ Such effects can be explained by prior findings of B[a]P-driven chromatin rearrangement and transcriptional blocking in male gametes, which can interfere with embryonic development.^31,37^ Both BPA and B[a]P have been associated with miscarriage, low birth weight, and other adverse pregnancy outcomes.^1,32,33^

Molecular interactions (Figure 5A and 5B) show that both BPA and B[a]P are linked, via curated CTD evidence, to interactions with the same genes that are most abundant in our developmental lethality set, and pathway enrichment (Figure 5C) indicates that both activate downstream coagulation-and implantation-related biology. This is directly in line with our earlier results at the network level (Figure 4) and lethality timing (Figure 3), where *F5*, *F2*, and *AURKB* were the most interconnected and most vulnerable in the second trimester, a critical window for placental development. Thus, Figures 4 and 5 are consistent with prior toxicogenomic and reproductive biology literature and also identify specific chemical pathways in genes that can influence RPL, suggesting a convergent mechanism of vascular or hemostatic compromise in early pregnancy failure.

These observations support a more explicit mechanistic model linking chemical exposure to recurrent pregnancy loss. At the uteroplacental interface, pregnancy depends on tightly regulated hemostasis, trophoblast invasion, and early embryonic competence.^19,20^ In this context, the BPA-and B[a]P-associated perturbations of *F2* and *F5* are consistent with dysregulated thrombin generation and fibrin formation, which could promote microvascular thrombosis, impaired placental perfusion, and implantation failure.^19,20^ The accompanying perturbation of *FOXD1* provides a complementary placental mechanism, as altered *FOXD1* expression could impair trophoblast differentiation and spiral artery remodeling, thereby worsening uteroplacental insufficiency.^32,21^ In parallel, the BPA-associated effects on *AURKB* suggest a possible route through meiotic spindle checkpoint disruption and chromosome mis-segregation, whereas methylation changes in *PADI6* are notable because *PADI6* is a maternal-effect gene required for oocyte competence and early embryonic development.^12,22–24^ These gene-level links suggest that BPA and B[a]P may converge on pregnancy loss through partially distinct but biologically coherent routes involving coagulation imbalance, defective placentation, and impaired early embryonic viability. Given the curated and hypothesis-generating nature of the CTD-derived interactions, these mechanisms should be interpreted as plausible models for experimental follow-up rather than direct evidence of causality.

However, some notable limitations persist. First, incomplete or missing gene annotations, especially in the Intolerome database, limit the full characterization of some genes and their roles in developmental disruption. Second, because CGA is anchored to an Intolerome-defined gene set, the resulting network reflects a prioritized subset of genes already implicated in developmental lethality rather than an unbiased survey of all human genes that may be influenced by environmental exposures. Third, the CTD database does not account for exposure heterogeneity, dose, duration, timing, co-exposures, emerging contaminants, or detailed exposure scenarios, which limits its ability to model real-world exposure conditions with sufficient granularity. Additionally, the absence of specific population-level features in our dataset, such as ancestry, geography, and individual exposure context, restricts the broader generalizability of our results.

Our findings also depend on curated literature in CTD and are therefore subject to both database and literature bias. Chemicals and genes that are more extensively studied, such as BPA and benzo[a]pyrene, may appear disproportionately connected in the network relative to less-studied exposures or less-characterized genes. In addition, curated chemical-gene interactions in CTD vary in evidentiary basis and mechanistic specificity, meaning that not all links should be interpreted as equivalent in biological strength or causal relevance. Consequently, network density or node degree should not be interpreted as a direct measure of biological specificity, causal importance, or real-world exposure burden.

Finally, the absence of experimental validation limits mechanistic interpretation. To enhance translational potential and strengthen the biological relevance of our findings, future research should incorporate dynamic databases of chemical exposure, examine populations with diverse backgrounds, and perform targeted validation of high-priority chemical-gene pairs in independent cohorts and biologically relevant experimental systems, including placental, trophoblast, embryo-relevant, and model-organism settings.

Despite these constraints, our findings point to clear opportunities for research translation. CGA can help researchers connect environmentally modifiable exposures to loss-of-function–intolerant genes, supporting etiologic hypotheses, and prioritizing follow-up studies across reproductive health outcomes.

Looking ahead, CGA offers a scalable and extensible framework for examining chemical-gene interactions beyond the specific case of RPL. It supports precision exposomics by allowing queries filtered by gene type (fetal or maternal), biological system, disruption type (loss-or gain-of-function), and timing of lethality. For example, users can identify placental genes expressed in the second trimester that are susceptible to environmental toxicants. Expanding CGA with additional datasets and real-world exposure profiles will further strengthen its value as a predictive toxicology and developmental risk assessment tool.

This exposomic analysis reveals that genes with LoF intolerance, particularly those active in metabolic, cardiovascular, and neurological systems, are disproportionately impacted by environmental chemical exposures. By integrating CTD and Intolerome datasets, we created a curated map of 928 vulnerable genes and ∼4000 chemical interactors, emphasizing genetic susceptibility during fetal and postnatal stages of development. The observed timing of lethality suggests latent, environmentally driven disruptions during critical physiological transitions, such as organogenesis and immune maturation. Exposomic profiling of five RPL-associated genes revealed conserved toxicant interactions, notably with bisphenol A and benzo[a]pyrene, which affected gene expression and methylation in coagulation and placental pathways. By integrating genomic intolerance and environmental interaction data, CGA provides a scalable, interactive tool for investigating and communicating clinically relevant, environmentally modifiable genetic risks.

## Resource Availability

### Lead contact

Requests for further information and resources should be directed to and will be fulfilled by the lead contact, Marina Sirota.

### Materials availability

This study did not generate new, unique reagents.

### Data and code availability

The datasets analyzed in this study are publicly available from the Intolerome and the Comparative Toxicogenomics Database (CTD). The curated and merged dataset, along with the source code for the Chemical-Gene Atlas (CGA), is openly accessible at https://github.com/SyedHassan20/cga-chem-gene-atlas. The CGA web application can be accessed at https://cgatlas.org/.

## Data Availability

All data produced in the present study are available upon reasonable request to the authors

https://rpldb.org/intolerome/

## Acknowledgements

The authors would like to thank Elizabeth Thomson of Peraton for her help, and Ana Maria Deluca, Grace Loll, and Edna Rodas of the UCSF Bakar Computational Health Sciences Institute for administrative support. Figures were created with BioRender.com.

This work was funded by the National Institutes of Health (NIH) Eunice Kennedy Shriver National Institute of Child Health and Human Development (NICHD) [R01 HD105256] and the March of Dimes Prematurity Research Center at UCSF [60982053-50185]. The funders played no role in the study design, data collection, analysis and interpretation of data, or the writing of this manuscript.

## Author Contributions

Conceptualization: M.S., G.D.S.C.; Data Curation: S.H.B., A.N., S.A.Y.; Formal Analysis: S.H.B.; Methodology: S.H.B., G.D.S.C.; Software: S.H.B., P.M.; Visualization: S.H.B., B.O., T.T.O.; Resources: M.S., A.R.; Investigation:; S.H.B., A.N.; Supervision: M.S., T.T.O., A.R.; Project Administration: T.T.O., M.S.; Writing-original draft: S.H.B., A.N.; Writing-review and editing: S.H.B., A.N., B.O., L.A., P.M., G.D.S.C., D.A.M., G.M.S., D.K.S., R.B.L., S.A.Y., T.T.O., A.R., M.S.; Funding Acquisition: M.S., A.R.

## Declaration of generative AI and AI-assisted technologies in the writing process

During the preparation of this work, the authors used OpenAI’s ChatGPT-4o and ChatGPT-5 models to improve readability and language, assist with code debugging, and support minor figure and symbol formatting tasks. After using these tools, the authors carefully reviewed and edited the content as needed and take full responsibility for the content of the published article.

## Methods

CGA utilizes a pre-merged dataset made from two public datasets: Intolerome, which provides data on genetic intolerance and gene-disease association (links between genetic variants and elevated risk of clinical outcomes such as developmental disorders or pregnancy loss), and CTD (Comparative Toxicogenomics Database, revision number 17996, downloaded November 2024), containing curated chemical-gene interactions (evidence that chemical exposures alter gene expression, activity, or function) and disease associations.^8,9,10^ The Intolerome dataset is publicly available at (https://rpldb.org/intolerome/), and CTD data were obtained from the download portal (https://ctdbase.org/downloads/). The merged CGA dataset and associated processing resources are available at: https://github.com/SyedHassan20/cga-chem-gene-atlas.

The merged dataset was constructed by matching Intolerome genes to CTD chemical-gene interaction records using OMIM Gene ID. Intolerome genes that did not map to CTD records through OMIM Gene ID were not retained in the merged dataset. The initial gene set comprised Intolerome genes associated with prenatal and postnatal disease phenotypes, genetic lethality, and developmental disorders (Figure 1). From CTD, we retained only curated chemical-gene interaction records involving genes from this Intolerome-derived set. As CTD includes curated interactions across multiple species, we restricted all chemical–gene interactions to *Homo sapiens* by applying organism-level filtering using the annotated organism field prior to merging with the Intolerome gene set (Figure 1). All curated human CTD chemical-gene interaction records linked to the retained Intolerome-derived genes were eligible for inclusion, regardless of the specific molecular interaction type annotated in CTD.

### Data Cleaning

The dataset underwent multiple preprocessing and cleaning steps before and after the merge. Before the merge, redundant white spaces, symbols, and non-informative placeholders were removed to create a more consistent and uniform dataset. Columns were restructured to ensure consistency between the pre-and post-merged datasets. Furthermore, the lethality timing data were reshaped from wide to long format for flexible downstream filtering and visualization. Finally, duplicate records were removed after harmonization by retaining one row per unique chemical-gene-interaction annotation combination (Figure 1). This step reduced redundant entries and improved interpretability in downstream network visualization and analysis.

### User Interface

We used HTML and CSS in R Shiny to structure and style the user interface for the web application. Dynamic filter controls were created to ensure a smooth filtering process. Four main filter dropdown menus are presented:

### 1. Gene Effect

Gene effect categorizes genes based on their roles in developmental stages, particularly fetal and neonatal outcomes. In this context, effect refers to the degree to which variation in a gene contributes to measurable risks, such as growth restriction, congenital malformations, or prenatal/neonatal lethality. By capturing these associations, the framework highlights how genetic variation influences developmental phenotypes and organizes genes into four categories: fetal, maternal, maternal–fetal, and unknown.

### 2. System Affected

This category organizes genes by the biological systems they affect, such as the cardiovascular, nervous, or metabolic systems. Therefore, it provides an opportunity for targeted investigation of system-specific disorders involving disrupted disease mechanisms and pathways.

### 3. Lethality Mode of Function

Lethality Mode of Function differentiates gene variants by their functional impact, primarily distinguishing loss-of-function and gain-of-function variants, with rare cases of duplication and deletion in our dataset. This distinction enables researchers to focus on genes that drive lethality and developmental anomalies, thereby devising therapeutic strategies and interventions.

### 4. Lethality Timing

Lethality Timing categorizes gene impacts by the developmental timing of lethality, spanning various trimesters through the postnatal period. This temporal categorization will allow researchers to target specific windows to their disease of interest.

These filters dynamically update downstream visualizations and tables. Any action taken by the users undergoes reactive selections implemented through Shiny’s reactive programming to ensure connectivity between different app components.

### Donut Chart Visualizations

The four filters are implemented using interactive donut charts to summarize the distributions of each filter category. These plots are also reactive upon user selection, and each click on a segment updates the network plots, diving deeper and narrower into the interactions between genes and chemicals (Figure 1).

### Network Plots

The second component of this app is a visualization of the chemical-gene interactions using the visNetwork package.^38^ Red nodes represent genes, green nodes represent chemicals, and edges represent the interaction. Node sizes scale proportionally to node degree (i.e., the number of connections), allowing for the prioritization of highly interconnected genes. We have included interactive features such as hover effects, nearest-neighbor highlighting, node selection via dropdown, and smooth transitions to explore different pathways and focus on specific genes or chemicals for each filter (Figure 1).

### Filtered Data Table

The last component of the app is a filtered table that supports global and column-specific filtering (Figure 1). In addition to core interaction features, the table includes OMIM-derived annotations such as disease name, inheritance pattern, system affected, and prenatal phenotype (e.g., neonatal death, structural malformations), enabling phenotype-level stratification of gene–chemical associations. The filtered table includes identifier-based hyperlinks to external records in CTD, NCBI Gene, OMIM, and PubMed, allowing users to navigate from app entries to the corresponding source records. The filtered dataset can also be exported directly from the app.

### Reactive Logic and Performance Optimization

Given the size of the dataset and the number of chemical-gene interactions, reactive programming principles ensure responsive, accurate data representation in response to user inputs. Apart from the reactive filters mentioned above, individual plotting and data-rendering functions were encapsulated in reactive expressions, ensuring consistent data synchronization across all visual components and tables.

We took additional steps to improve performance, considering the computational complexity of the task. We leveraged asynchronous rendering with the Future and Promise libraries to manage computationally intensive tasks, such as network plotting, without causing interruptions.^39,40^ Visual feedback in the form of loader indicators was provided during intensive calculations to improve the user experience. Lastly, we increased the maximum allowable global memory size to 600 MB when running multisession in R to prevent memory limit errors during parallel processing.

### Deployment and Accessibility

The CGA app was developed and extensively tested in an R environment (version 4.3.2). Complete source code and comprehensive documentation are publicly accessible through GitHub, providing reproducibility, transparency, and encouraging community-driven enhancements: https://github.com/SyedHassan20/cga-chem-gene-atlas.

### Reactome Pathway**-**Enrichment Workflow

For each toxicant, we filtered the interaction table for BPA or B[a]P, converted the resulting non-redundant gene symbols to Entrez IDs with bitr (org.Hs.eg.db), and ran enrichPathway (ReactomePA/clusterProfiler v4.8.3) against the default human Reactome universe using a one-tailed hypergeometric test; Benjamini-Hochberg-adjusted *p-value* < 0.10.^41,42^ This p-value is chosen to retain power for small, exploratory gene sets while remaining stricter than the package’s 20 % default, which was considered significant.

## Graphical Abstract. Integration of Intolerome Genes with Chemical-Gene Interaction Data to Construct a Chemical-Gene Network.

Input Data is an initial list of 934 Intolerome genes, filtered for *Homo Sapiens*, and merged with CTD (Comparative Toxicogenomics Database) records containing 28,015 genes, 10,809 unique chemicals, and 1,221,061 chemical-gene interactions. Processing and Integration of the system provides annotations for fetal or maternal gene effects and unknown effects with specific lethal mechanisms, such as DNA-disruptive nucleotide deletions, and developmental period information ranging from the first trimester into postnatal stages. The Chemical-Gene Network is a chemical-gene interaction network constructed, comprising 928 genes and 4,110 chemicals, yielding 73,256 unique chemical-gene interactions. This network provides a framework for investigating the impact of environmental exposures on vulnerable developmental windows via chemical-gene interactions.

